# Is convalescent plasma futile in COVID-19? A Bayesian re-analysis of the RECOVERY randomised controlled trial

**DOI:** 10.1101/2021.04.01.21254679

**Authors:** FW Hamilton, TC Lee, DT Arnold, R Lilford, K Hemming

## Abstract

**Introduction:** Randomised trials are generally performed from a frequentist perspective reporting point estimates and 95% confidence intervals. This approach can confuse “evidence of no effect” with “no evidence of an effect” and does not allow for contextual knowledge. The RECOVERY trial evaluated convalescent plasma for patients hospitalised with COVID-19, the interaction test for the primary outcome was not statistically significant, and the trial concluded no evidence of an effect. From the clinical immunology perspective, there is strong justification to expect differential responses to convalescent plasma in patients who already have their own antibodies to SARS-CoV2 (seropositive) versus those who do not (seronegative).

**Methods:** Outcome data was extracted from the RECOVERY trial both overall and for seronegative participants. A Bayesian re-analysis with a wide variety of priors (vague, optimistic, skeptical and pessimistic) was performed calculating the posterior probability for both any benefit or a modest benefit (number needed to treat of 100).

**Results:** Across all patients, when analysed with a vague prior the likelihood of any benefit or a modest benefit was estimated to be 64% and 18% respectively. In contrast, in the seronegative subgroup, the likelihood of any benefit or a modest benefit was estimated to be 90% and 74%. Results were broadly consistent across all prior distributions.

**Conclusion:** Performing clinical trials during a pandemic is challenging, and RECOVERY has provided high quality evidence for numerous therapies. However, the use of frequentist hypothesis testing in this trial has led to the trialists and governing bodies to conclude a strong evidence of no effect. Based on this trial, and other prior knowledge there remains a strong probability that convalescent plasma provides at least a modest benefit in seronegative patients.

## Introduction

Convalescent plasma (CP) – blood components from patients recovered from an infection - has been used for more than a century to treat infections, with widespread use in the 1920’s and 30’s for pneumococcal infections and scarlet fever, before falling out of favour with the development of antibiotics.^1^ The principle is that of ‘passive immunisation’ – passing antibodies from those recovered from infection to those naïve to it, and therefore providing a degree of protection from that specific agent.^2^ It is therefore unsurprising that interest in the use of CP to prevent and treat COVID-19 has been widespread.^1^ Unfortunately, despite best efforts, most of this usage has occurred outside of randomised controlled trials (RCT) with >100,000 doses given in the US alone. ^3^

Fortunately, the RECOVERY collaborative group have recently reported the largest randomised control trial of CP in hospitalised patients with COVID-19^4^. The authors conclude that CP provided no benefit, with mortality equal in both arms: 1398 (24%) of 5795 patients allocated to convalescent plasma and 1408 (24%) of 5763 patients allocated to usual care died within 28 days (rate ratio [RR] 1.00; 95% confidence interval [CI] 0.93 to 1.07; p=0.93). They also conclude there was no difference across pre-specified subgroups including those with detectable SARS-CoV-2 antibody tests at the time of randomisation (seropositive group), 19% versus 18% (RR 1.05; 95% CI, 0.93 to 1.19) and seronegative patients, 32% versus 34% (RR 0.94; 95% CI, 0.84 to 1.06); with test for interaction p=0.21. In particular, they note, on the advice of the Drug Safety and Monitoring Committee (DMC), that: “*there was no convincing evidence that further recruitment would provide conclusive proof of worthwhile mortality benefit either overall or in any pre-specified subgroup*.” In the United Kingdom, this data has been taken by the regulator as strong evidence of a null effect, leading the Medicines Health Regulatory Authority (MHRA), the UK medicines regulator, to recommend against the use of CP in patients hospitalised with COVID-19, effectively removing the therapy in the National Health Service (NHS). ^5^

Before accepting that CP is ineffective in hospitalized patients, it is important to recognise the clear distinction between patients who are seronegative (have no detectable antibodies to SARS-CoV-2 at admission), and seropositive (those that do). The therapeutic mechanism of convalescent plasma and monoclonal antibody (e.g. REGN-COV2) treatments is by passive immunisation – the gifting of antibodies. It is not surprising to think that the greatest (or any) benefit of CP would only occur in patients who are seronegative, or conversely, that there will be little to no benefit in giving antibodies to those who already have antibodies. Previous literature from SARS supports this^6,7^, as well as data clearly identifying a protective effect of monoclonal antibodies (manufactured antibodies, rather than donated) in early COVID-19 trials, with much weaker effects in hospitalised patients later in the disease course.^8–11^ Immunological data and cases of persistent infection show that failure of an early antibody response is associated with both severe disease, and in patients without any antibodies, the risk of persistent disease.^12,13^Others have also argued that seropositivity is a reason for failure of convalescent plasma.^14^

On that background, it is logical to analyse the data from patients who are seronegative (hypothesed more likely to benefit) separately from those who are seropositive (hypothesed less likely to benefit). Although subgroup analyses can be complicated by chance imbalances, lower power, and issues of multiple testing, they are appropriate to generate hypotheses and could be used in support of the argument of not disregarding CP as a potential treatment too soon.^15,16^ Moreover, conflating absence of evidence for a small effect with evidence of no effect further risks discarding a therapy which could still have a meaningful benefit. We therefore sought to undertake a Bayesian re-analysis to estimate the probability of (a) any benefit and (b) a modest benefit (which we define as equivalent to a number needed to treat of at most 100) using both the RECOVERY data overall and in the seronegative subgroup for whom the prior probability of success is hypothesized to be higher.

## Methods

We extracted the intention to treat results from the RECOVERY trial both overall and for the seronegative subgroup. Using STATA version 16 (Statacorp, College Station, TX) and the function bayes we calculated the posterior probabilities of (a) any benefit (OR <1) and (b) a modest but clinically important benefit estimated as absolute risk difference of at least 0.5% (number needed to treat <=200) or 1% (number needed to treat <=100). As suggested by a recent review on conducting Bayesian re-analysis in COVID-19,^17^ we chose four probability assumptions to account for varying prior views: vague (no information : mean risk difference (RD) 0, SD 10,000), optimistic (10% risk of harm: mean RD 0.01, SD 0.007), sceptical (tightly around the null: mean RD 0, SD 0.007), and pessimistic (10% chance of benefit (mean RD –0.005, SD 0.0036). Posterior probabilities were computed from binomial regression models.

We also used the R Shiny app Bayesian re-analysis of Clinical Trials^18^ (based on methodology from Wijeysundera et al.^19^) to produce heatmaps showing the prior probability assumptions required to generate posterior probabilities of any benefit for both the whole study and the seronegative arm. All code used to generate these figures is available in the appendix.

## Results

Table 1 presents the posterior probabilities of benefit for each prior (optimistic, skeptical, and pessimistic) for any benefit, a risk difference of 0.5% (equivalent to a number needed to treat (NNT) of 200) and a risk difference of 1% (equivalent to a NNT of 100).

**Table 1:**
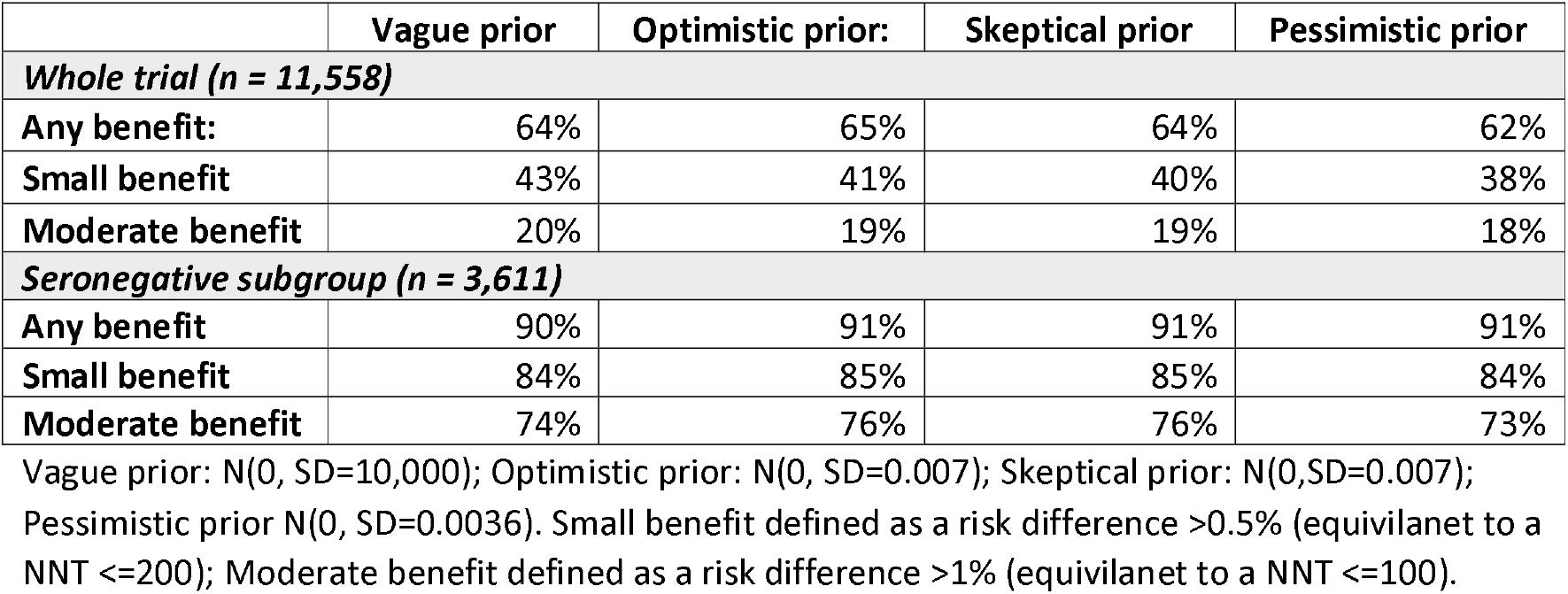
Estimated posterior probabilities of benefit for a variety of prior assumptions

|  | Vague prior | Optimistic prior: | Skeptical prior | Pessimistic prior |
| --- | --- | --- | --- | --- |
| **Whole trial (n = 11,558)** |  |  |  |  |
| **Any benefit:** | 64% | 65% | 64% | 62% |
| **Small benefit** | 43% | 41% | 40% | 38% |
| **Moderate benefit** | 20% | 19% | 19% | 18% |
| **Seronegative subgroup (n = 3,611)** |  |  |  |  |
| **Any benefit** | 90% | 91% | 91% | 91% |
| **Small benefit** | 84% | 85% | 85% | 84% |
| **Moderate benefit** | 74% | 76% | 76% | 73% |

Vague prior: $N(0, SD=10,000)$ ; Optimistic prior: $N(0, SD=0.007)$ ; Skeptical prior: $N(0, SD=0.007)$ ; Pessimistic prior $N(0, SD=0.0036)$ . Small benefit defined as a risk difference $>0.5\%$ (equivalent to a NNT $\leq 200$ ); Moderate benefit defined as a risk difference $>1\%$ (equivalent to a NNT $\leq 100$ ).

Across the whole trial population, the estimated chance of benefit is around 65%, with little difference depending on prior assumptions. The posterior probability of a risk difference of >1% (preferring treatment arm) is around 19 % across all prior assumptions. The associated heatmaps of posterior probability as a function of the initial prior assumptions are presented in figure 1 (presented on the odds ratio scale).

**Figure 1:**
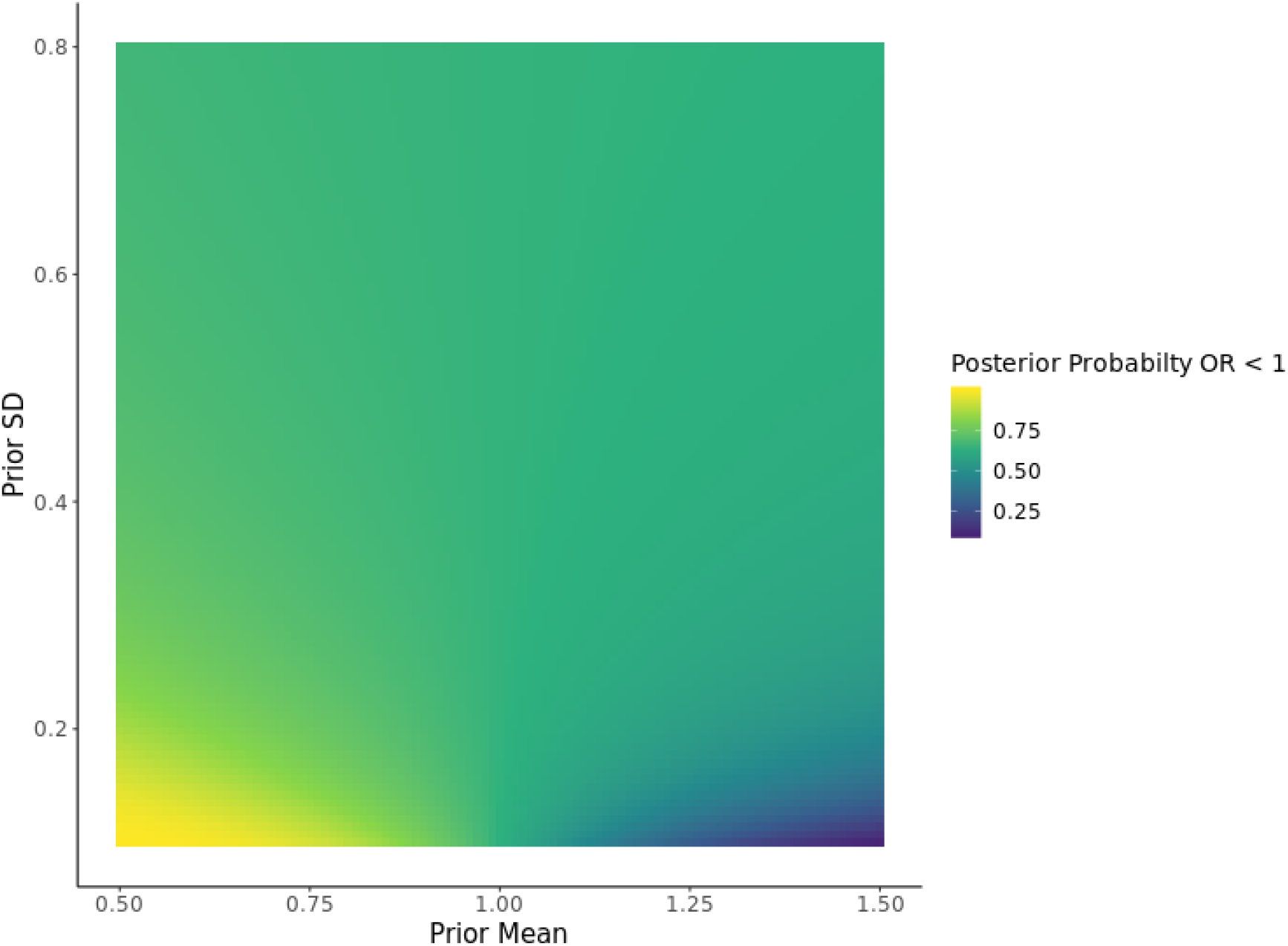
Heatmap of posterior probability of any benefit (OR of <1) of convalescent plasma for all trial participants.

In the seronegative subgroup, the estimated likelihood of any benefit is greater, at around 90%, across all prior assumptions. The estimated chance of a risk difference of >1% is also high (more than 73% across all three priors), and varied little between prior assumptions. The associated heatmaps of posterior probability as a function of the initial prior assumptions are presented in figure 2.

**Figure 2:**
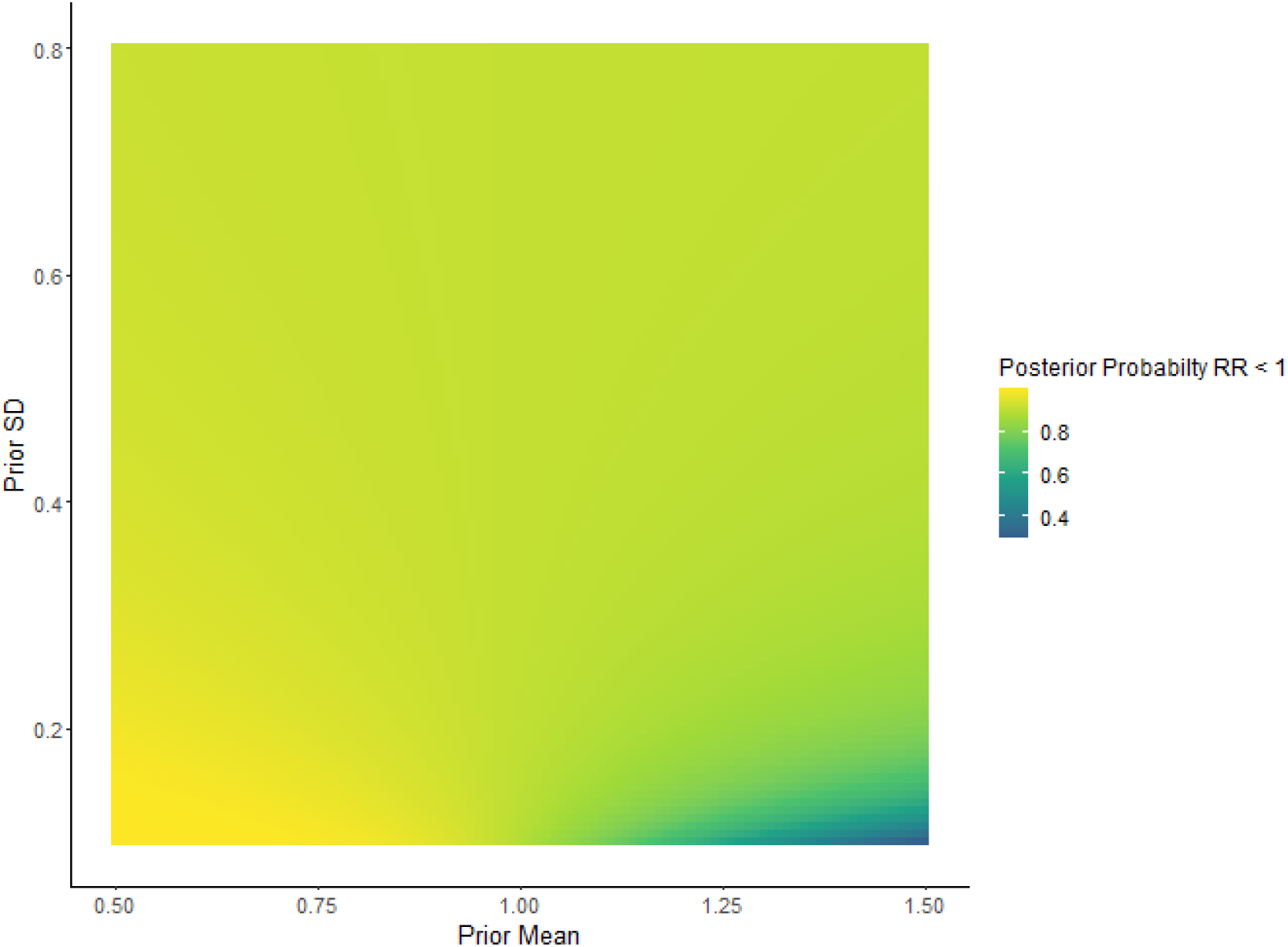
Heatmap of posterior probability of any benefit (OR of <1) of convalescent plasma for all the seronegative participants only

## Discussion

The RECOVERY trial has been a paradigm in a rapid, pragmatic, approach to trialling new therapies in a pandemic. Good practice requires a firm, pre-specified analysis plan with clear, pre-defined subgroup analysis. ^20^ However, the conclusions drawn by the authors and Medicines Health Regulatory Authority (MHRA) with respect to convalescent plasma risks conflating absence of evidence of a small effect with evidence that there is no benefit. In fact, the data presented are consistent with a small probability (>15%) of an effect of with an NNT of 100 across the whole trial, with an even higher probability (>73%) if analysing the seronegative arm separately.

Many clinicians, patients and their families might consider benefits in the region of 1 life saved for every 100 or 200 people treated as meaningful benefits; and thus, there remains an important chance that there could remain a clinically important benefit among patients who are seronegative. Benefits of this magnitude are not only important to patients but have potentially large public health effects when considering the scale of the pandemic. However, very large trials are needed to detect such small differences, often so large that they are financially or logistically not feasible. Previous trials have been small and underpowered, with a recent meta-analysis identifying less than 2,000 patients across all RCT’s prior to RECOVERY.^21^ Only one previous trial of high titre CP has reported ^22^data based on antibody status ; with 34/105 deaths in the seronegative placebo arm and 65/228 deaths in the CP arm, a RR of death with CP 1.12 (95%CI: 0.51-2.43), providing little information. In contrast the largest observational study in hospitalised patients (n = 3,082), a US registry study (published after RECOVERY), identified a lower risk of death in patients transfused early with higher levels of SARS-CoV-2 IgG antibody, providing further support for our prior estimates of benefit. ^23^

Whilst all trials have to be conducted within the context of these pragmatic constraints, it is nonetheless important to differentiate between those treatments which we know are very unlikely to work, and those which we are unable to make conclusions.

We recognize that there may have been chance imbalances in age or comorbidity within the seronegative subgroup of patients since randomization was not stratified on serostatus.^4^ Nonetheless, we feel it is appropriate to analyse this group in the context of the other available data (that early monoclonal antibody is most effective in seronegative patients, for example), and the increasing recognition that serostatus is likely a key predictor of response to therapy. Therefore, in this context, the focus on this subgroup is an appropriate response to greater understanding of the causal path by which CP might be effective. An analysis which adjusted for demographic differences between the CP and usual care groups could provide more precise estimates as could pooling patient data from all other high titre trials where serostatus was determined.

Additionally, although we focused our analysis entirely on the primary outcome to avoid “cherry picking” data, it is also important to note that both secondary outcomes (discharge home by day 28, and invasive mechanical ventilation or death) did show heterogeneity with respect to serostatus (p = 0.003 and p = 0.01), with both actually showing benefit in the CP arm using frequentist null hypothesis significance testing.

Given the challenges and costs of implementing CP in seronegative patients (requirement for rapid testing, blood typing, alongside an efficient blood transfusion laboratory), we recognise that others may disagree on what is a “clinically meaningful” effect size. Regardless, the important point is that the RECOVERY trial is leaves residual uncertainty for the seronegative group with a reasonable likelihood of benefit that some would consider important.

## Conclusions

The RECOVERY trial for CP reported no benefit. Recognising the changing literature since the trial started and using a variety of priors, we suggest the reporting of no effect may be premature. It remains plausible that CP has a small, but clinically important effect on mortality in those who have not already developed an antibody response. It is clear that any effect is likely small, but we would argue clinicians, scientists, and government agencies to review all trial data in totality, rather than regarding the null result as fixed.

## Supporting information

Stata Do file (can open in txt editor)

## Data Availability

All code is available in the appendix. All raw data is available in the RECOVERY pre-print

