## Supplementary material for "Is convalescent plasma futile in COVID-19? A Bayesian re-analysis of the RECOVERY randomised controlled trial": Stata Do file (can open in txt editor): bayesian_submission.do

```
clear
set seed 26032021
**********************************************
*********Seronegative population**************
*sero-negative 
local e_t=626
local e_c=549
local n_t=1982
local n_c=1629
**********************************************
**********************************************

**********************************************
**********************************************
*All population ******************************
**********************************************
**********************************************
*local e_t=1398
*local e_c=1408
*local n_t=5795
*local n_c=5763
*Generating individual-level data
local tss=`n_t' + `n_c'
di `n_c'
set obs `tss'
qui gen id = _n
qui gen Treat=0
qui replace Treat=1 if id<=`n_t'
qui gen Outcome=0
qui replace Outcome=1 if id<=`e_t'
qui replace Outcome=1 if id<=`n_t'+`e_c' & id>`n_t'
*Performing simple frequentist analysis 
cs Outcome Treat
**************************
*Bayes analysis Risk difference scale (used to compute NNT)
**************************
mat results_rd = J(12,3,.)
mat colnames results_rd = Post_prob SE MSCE
mat rownames results_rd = Anybenefit_vague NNT100_vague NNT200_vague AnyBenefit_optimistic NNT100_optimistic NNT200_optimistic AnyBenefit_pessimistic NNT100_pessimistic NNT200_pessimistic AnyBenefit_skeptical NNT100_skeptical NNT200_skeptical
mat results_or = J(4,3,.)
mat colnames results_or = Post_prob SE MSCE
mat rownames results_or =  AnyBenefit_vague AnyBenefit_optimistic AnyBenefit_pessimistic AnyBenefit_skeptical 
*Prior: off the shelf vague (stata default) mean 0 variance 10,000
bayes, rseed(29032021): binreg Outcome Treat, rd
bayestest interval {Outcome:Treat}, upper(0) /*** any benefit ***/
qui: return list
mat V=r(summary)
mat results_rd[1,1]=V[1,1]
mat results_rd[1,2]=V[1,2]
mat results_rd[1,3]=V[1,3]
bayestest interval {Outcome:Treat}, upper(-0.01) /*** NNT of 100 ***/
qui: return list
mat V=r(summary)
mat results_rd[2,1]=V[1,1]
mat results_rd[2,2]=V[1,2]
mat results_rd[2,3]=V[1,3]
bayestest interval {Outcome:Treat}, upper(-0.005) /***NNT of 200  ***/
qui: return list
mat V=r(summary)
mat results_rd[3,1]=V[1,1]
mat results_rd[3,2]=V[1,2]
mat results_rd[3,3]=V[1,3]

*Prior: optimistic 
*Informative prior set mean at NNT 100 
*SD such that 10% probanility that RD>0
*p(z> 1.38)=0.0985~0.1 (1.38 SDs from the mean) 
*Solve 1.38=(0+0.01)/SD; gives SD=0.01/1.38=0.007
*twoway function y = normalden(x, -0.01,0.007), range(-0.05 0.05)
bayes, rseed(29032021) prior({Outcome:Treat}, normal(-0.01,0.007)) : binreg Outcome Treat, rd
bayestest interval {Outcome:Treat}, upper(0) /*** any benefit ***/
qui: return list
mat V=r(summary)
mat results_rd[4,1]=V[1,1]
mat results_rd[4,2]=V[1,2]
mat results_rd[4,3]=V[1,3]
bayestest interval {Outcome:Treat}, upper(-0.01) /*** NNT of 100 ***/
qui: return list
mat V=r(summary)
mat results_rd[5,1]=V[1,1]
mat results_rd[5,2]=V[1,2]
mat results_rd[5,3]=V[1,3]
bayestest interval {Outcome:Treat}, upper(-0.005) /***NNT of 200  ***/
qui: return list
mat V=r(summary)
mat results_rd[6,1]=V[1,1]
mat results_rd[6,2]=V[1,2]
mat results_rd[6,3]=V[1,3]


*Prior: pessimistic 
*Informative prior set mean at NNH 200
*SD such that 10% probanility that RD<0
*For SD solve SD=0.005/1.38=0.0036
*twoway function y = normalden(x, 0.005,0.0036), range(-0.05 0.05)
bayes, rseed(29032021) prior({Outcome:Treat}, normal(0.005,0.0036)) : binreg Outcome Treat, rd
bayestest interval {Outcome:Treat}, upper(0) /*** any benefit ***/
qui: return list
mat V=r(summary)
mat results_rd[7,1]=V[1,1]
mat results_rd[7,2]=V[1,2]
mat results_rd[7,3]=V[1,3]
bayestest interval {Outcome:Treat}, upper(-0.01) /*** NNT of 100 ***/
qui: return list
mat V=r(summary)
mat results_rd[8,1]=V[1,1]
mat results_rd[8,2]=V[1,2]
mat results_rd[8,3]=V[1,3]
bayestest interval {Outcome:Treat}, upper(-0.005) /***NNT of 200  ***/
qui: return list
mat V=r(summary)
mat results_rd[9,1]=V[1,1]
mat results_rd[9,2]=V[1,2]
mat results_rd[9,3]=V[1,3]
*Prior: Skeptical 
*Informative prior set mean at 0
*For SD 0.007 quite tight around null of no effect 
*twoway function y = normalden(x, 0.00,0.0007), range(-0.05 0.05)
bayes, rseed(29032021)  prior({Outcome:Treat}, normal(0,0.007)) : binreg Outcome Treat, rd
bayestest interval {Outcome:Treat}, upper(0) /*** any benefit ***/
qui: return list
mat V=r(summary)
mat results_rd[10,1]=V[1,1]
mat results_rd[10,2]=V[1,2]
mat results_rd[10,3]=V[1,3]
bayestest interval {Outcome:Treat}, upper(-0.01) /*** NNT of 100 ***/
qui: return list
mat V=r(summary)
mat results_rd[11,1]=V[1,1]
mat results_rd[11,2]=V[1,2]
mat results_rd[11,3]=V[1,3]
bayestest interval {Outcome:Treat}, upper(-0.005) /***NNT of 200  ***/
qui: return list
mat V=r(summary)
mat results_rd[12,1]=V[1,1]
mat results_rd[12,2]=V[1,2]
mat results_rd[12,3]=V[1,3]
*List results 
mat list results_rd

*Plot overlaying all priors 
twoway (function y = normalden(x, -0.01,0.007), range(-0.05 0.05)) ///
(function y = normalden(x, 0.005,0.0036), range(-0.05 0.05)) ///
(function y = normalden(x, 0.00,0.007), range(-0.05 0.05)) ///
(function y = normalden(x, 0.00,1000), range(-0.05 0.05)), ///
legend(order(1 "Optimistic" 2 "Pessimistic" 3 "Sceptical" 4 "Vague" )) 
**************************
*Bayes analysis OR scale (used to compute on relative scale)
**************************
*For priors following: Zampieri FG, Casey JD, Shankar-Hari M, Harrell FE Jr, Harhay MO. Using Bayesian Methods to Augment the Interpretation of Critical *Care Trials. An Overview of Theory and Example Reanalysis of the Alveolar Recruitment for Acute Respiratory Distress Syndrome Trial. Am J Respir Crit Care *Med. 2021 Mar 1;203(5):543-552. doi: 10.1164/rccm.202006-2381CP. PMID: 33270526; PMCID: PMC7924582.
*Used moderate priors (as defined below and in above paper, middle plots)
*Prior: off the shelf vague (stata default)
bayes, rseed(29032021) : binreg Outcome Treat, or
bayestest interval {Outcome:Treat}, upper(0) /*** any benefit ***/
qui: return list
mat V=r(summary)
mat results_or[1,1]=V[1,1]
mat results_or[1,2]=V[1,2]
mat results_or[1,3]=V[1,3]
*Prior: optimistic moderate
*mean lnOR=-0.41 equivalent to OR 0.64
*SD 0.3 means there is a 8% probability that the OR >1 (so very optimistic); called "moderate strengh"
bayes, rseed(29032021) prior({Outcome:Treat}, normal(-0.41,0.4)) : binreg Outcome Treat, or
bayestest interval {Outcome:Treat}, upper(0) /*** any benefit ***/
qui: return list
mat V=r(summary)
mat results_or[2,1]=V[1,1]
mat results_or[2,2]=V[1,2]
mat results_or[2,3]=V[1,3]

*Prior: pessimistic moderate
*mean lnOR=0.41 equivalent to OR 1.55
*SD 0.7 means there is a 27% probability that OR <1 (so very pessimistic) called "moderate strengh"
bayes, rseed(29032021) prior({Outcome:Treat}, normal(0.41,0.6)) : binreg Outcome Treat, or
bayestest interval {Outcome:Treat}, upper(0) /*** any benefit ***/
qui: return list
mat V=r(summary)
mat results_or[3,1]=V[1,1]
mat results_or[3,2]=V[1,2]
mat results_or[3,3]=V[1,3]
*Prior: skeptical 
*Mean centred at zero
*SD at 0.355
bayes, rseed(29032021)  prior({Outcome:Treat}, normal(0,0.355)) : binreg Outcome Treat, or
bayestest interval {Outcome:Treat}, upper(0) /*** any benefit ***/
qui: return list
mat V=r(summary)
mat results_or[4,1]=V[1,1]
mat results_or[4,2]=V[1,2]
mat results_or[4,3]=V[1,3]

mat list results_or

clear
```
